# Genetic Signal Augmentation of Childhood-Onset and Treatment-Resistant Major Depression Reveals Distinct Biological Disorders

**DOI:** 10.64898/2026.03.02.26347449

**Authors:** Jeremy M. Lawrence, Sophie Breunig, Lukas S. Schaffer, Alexander Sheppard, Katerina Zorina-Lichtenwalter, Andrew D. Grotzinger

## Abstract

Major depression (MD) is a disorder class that exhibits substantial phenotypic and clinical heterogeneity, yet many large-scale molecular genetic investigations treat MD as a unitary outcome. Here, we applied Genomic Structural Equation Modeling (Genomic SEM) to characterize the genetic variation in two clinically relevant MD subtypes, childhood-onset (child-onset) and treatment-resistant MD, that are independent of the field-standard GWAS of MD in all its forms. In addition, we fit a complementary “boosting” model that leveraged shared signal across the subtype and general MD GWAS to increase power for subtype biological discovery. At the genome-wide level, more than half of the common-variant liability for child-onset and treatment-resistant MD was unique relative to the general MD GWAS, indicating substantial subtype-specific genetic architecture. Unique components of both subtypes showed robust associations with genetic liability for schizophrenia and bipolar disorder, and the child-onset specific component exhibited genome-wide overlap with early developmental outcomes, including autism spectrum disorder and childhood intelligence. Transcriptome-wide analyses implicated upregulation of *SMIM19* in liability specific to child-onset MD, while stratified functional enrichment highlighted gene sets involved in limbic and frontal brain systems for the boosted child-onset component. Together, these findings demonstrate that MD contains biologically distinct subtypes that exhibit etiological divergences more akin to separate disorders than subtypes within a concrete diagnostic framework. We find that stratifying MD by biologically distinguishable subtypes may be crucial for enhancing biological discovery and elucidating etiological pathways in molecular genetic studies of depression.

## Introduction

Major Depression (MD), characterized by markedly diminished affect and anhedonia^1^, is one of the most significant global health challenges, with Global Burden of Disease studies consistently ranking MD among the leading causes of global disability^2,3^. Pharmacological and psychotherapeutic treatment of MD is heterogeneous^4–8^, and interventions have variable efficacy, with a large proportion of treatment-resistant cases^9^. There is further variability at the nosological level. Under current diagnostic frameworks^1,10^, numerous symptom combinations are considered criteria for MD. For example, hypersomnia, insomnia, weight gain, and weight loss - traits with an extant literature describing distinct risk architectures^11–14^ - are all criteria for a diagnosis. Phenotypic variability embedded in the MD diagnosis reflects the assumption that, despite this variability, a common etiology underlies the different clinical presentations. We test this assumption here.

Large-scale genome-wide association studies (GWAS) of MD by the Psychiatric Genomics Consortium (PGC) have established a highly polygenic architecture and amassed among the largest case observations in psychiatric genetics for any disorder^15,16^. However, as case observations have increased, the population variance, *SNP*□based heritability (*h*^*2*^_*SNP*_), explained by common genetic variants, has failed to yield the gains observed in other mood disorders^17^. Although this trend could reflect a closer approximation to a ground truth that MD is less heritable, it may also indicate the conflation of multiple, etiologically distinct subtypes into a unitary diagnostic construct, resulting in a diffuse and attenuated overall genetic signal. This is because regions with a strong effect on only one subtype will have their signal diluted towards the null when averaged with the signal for other subtypes. Indeed, GWAS of MD sub-presentations (e.g., atypical, early-onset, treatment resistant) consistently observe higher *h*^*2*^_*SNP*_ at lower case observations^18,19^, with genetic correlations (*r*_*g*_) suggesting distinct genetic architectures subsumed within the general diagnostic construct^18^. The genetic architecture observed in large-scale investigations of MD and subtypes gives credence to the heterogeneity dilution hypothesis for conventional GWAS of general MD.

To deconstruct this heterogeneity and isolate more tractable genetic signals, we focus on two clinically relevant subtypes hypothesized in the extant literature to be particularly etiologically distinct and observed in this study to have significant unique variability relative to the general MD construct. From a developmental perspective, we analyzed childhood-onset MD (child-onset), a presentation with higher familial loading and a more severe prognosis^20–22^. Targeting this subtype aims to capture a potent genetic signal that, by comparison, is less confounded by what are likely more diverse environmental risks contributing to the later onset of depression in adulthood. From a severity perspective, we employed an extreme phenotype design by evaluating treatment-resistant MD^9,23^, enabling a direct test of whether heightened clinical severity reflects a greater accumulation of a common MD liability, as opposed to higher burden risk alleles.

We apply Genomic Structural Equation Modeling^24^ (Genomic SEM) to disambiguate the shared and unique genetic architecture when comparing these two MD sub-presentations to general MD. We conducted the first publicly available GWAS of child-onset MD, restricting cases to onset ≤ 18, alongside an existing GWAS of treatment-resistant MD^19^ to partition their genetic signals relative to the general PGC MD GWAS. Our approach is twofold: first, we used Cholesky decomposition to isolate the genetic signal unique to each subtype that is independent of general MD. Second, we employed a complementary model to boost statistical power for discovering subtype signals contained within the general MD GWAS. This dual framework allows us to gain novel biological insights across multiple levels of analysis, from patterns of genetic risk sharing with clinically relevant traits to tissue-specific gene expression and functional pathways that characterize these two distinct MD subtypes.

## Methods

### MD Subtype Selection and GWAS

We created an initial list of theoretically relevant MD subtypes based on the existing literature (child-onset, treatment-resistant, postpartum, and atypical), noting that we focus our primary results on child-onset and treatment-resistant due to the greater levels of unique genetic signal (described below). We present case-control information for all MD subtypes in Supplementary Table 1. This included GWAS summary statistics for treatment-resistant MD from the Swedish PREFECT study^19^. We also ran our own GWAS in the UK Biobank (UKB) for child-onset, postpartum, and atypical MD using SAIGE v1.30^25^. We restricted cases and controls based on the completion of the online Mental Health Questionnaire (MHQ). We apply this restriction due to existing literature documenting demographic and health-related differences between MHQ responders and non-responders^26^. Given these differences, prior work on the genetics of MD subtypes that utilized the MHQ for MD cases and individuals who did not take the MHQ for controls^18^ risks identifying pathways that characterize the subset of the UKB that took the MHQ, rather than true subtype signal.

We defined broad MD as a composite case status that classified participants as cases if they met any of several depression definitions, CIDI-based current or lifetime, endorsement of cardinal symptoms, hospital-record ICD-coded depressive disorder, probable depression, or self-reported depression. Child-onset MD cases were defined using this broad MD definition and retrospective self-report of age at first depressive episode between ages 2 and 18 years. We defined atypical MD as broad MD cases who additionally endorsed hypersomnia and weight gain. Postpartum MD cases included broadly-defined MD cases with either self-report or hospital records corresponding to depression occurring in relation to pregnancy/childbirth.

We applied FUMA v1.5.2^27^ to identify significant genomic risk loci, using a significance threshold of *p* < 5 × 10^−8^. We defined a genetic locus as genome-wide significant SNPs not in linkage disequilibrium (LD) with other significant SNPs at a threshold of *R*^2^ > 0.1 from the LD structure contained within the 1000 Genomes Phase 3 European ancestry reference panel. Genomic loci within 250 kilobases (kb) were merged into a single locus, as proximal SNPs may not have surpassed the LD threshold due to a mismatch between the GWAS samples and the reference panel.

### Multivariable LDSC

We then estimated genetic correlations with multivariable Linkage Disequilibrium Score Regression (LDSC)^28^ implemented in Genomic SEM^24^. Prior to running LDSC, a standard set of QC procedures was applied to the GWAS data, including filtering to HapMap3 SNPs and removing SNPs with a minor allele frequency (MAF) < 0.01 and imputation score (INFO) < 0.9. Multivariable LDSC estimates the genetic covariance and sampling covariance matrices across included traits. The genetic covariance matrix contains SNP-based heritability estimates on the diagonal and genetic covariances on the off-diagonal. For binary traits (e.g., the case/control GWAS of MD and its subtypes), these estimates were converted to the more interpretable liability scale using the population prevalence from the corresponding publication and the sum of effective sample size across cohorts contributing to the GWAS^29^. The sampling covariance matrix contains squared standard errors on the diagonal (the sampling variances) and sampling covariances on the off-diagonal, which index sampling dependencies arising from overlapping participant samples in the included GWAS data. We used 1000 Genomes Phase 3 European LD weights to estimate the regression model, excluding the major histocompatibility complex because its complex LD structure biases derived estimates.

### Genomic SEM

After estimating the genetic covariance and sampling covariance matrix, we applied Genomic SEM^24^ to fit a lower-triangle Cholesky decomposition model to the genetic overlap between general MD and each subtype examined. We began by regressing the general MD indicator on a single latent variable (*Dep*) and the subtype indicator on both *Dep* and a second, orthogonal latent variable (*uDep*). By fixing the residual variances of both the general and subtype MD GWAS to zero, we ensured that these two latent factors accounted for the entirety of the common variant genetic variance. This Cholesky decomposition specification enables us to distinguish the genetic signal shared with each subtype and general MD (*Dep*) from the genetic signal unique to each subtype (*uDep)*, effectively creating orthogonal variance components. We note here that the *Dep* factor reflects the total common variant signal in general MD including, but not restricted to the signal that is shared with the MD subtype included in each model.

We then pruned sub-presentations with non-significant unique variance components (*p >* .05). The two subtypes that surpassed these criteria were child-onset and treatment-resistant (ECT-treated) MD. Although atypical (*h*^*2*^_*SNP*_ *Z* = 6.81) and postpartum (*h*^*2*^_*SNP*_ *Z* = 4.80) MD had nonsignificant unique variance components, these estimates were substantial (27-41%), and their lack of significance was likely a function of reduced statistical power inherent in variance partitioning.

### External Trait Selection

We collected 16 publicly available GWAS summary statistics for psychiatrically relevant outcomes across cognitive, interpersonal, physical health, and psychiatric domains. All external traits had SNP-based heritability (*h*^*2*^_*SNP*_ ) Z-statistics > four, which reflects the threshold suggested by the original LDSC developers for producing interpretable estimates of genetic overlap^28^. These 16 traits were subsequently modeled alongside the unique and shared components of each MD subtype within Genomic SEM.

### Biological Annotation

We applied Transcriptome-wide SEM (T-SEM)^30^ and Stratified Genomic SEM^31^ to identify tissue-specific gene expression and differential enrichment for signal unique to child-onset and treatment-resistant MD. When underpowered to detect biological annotations for each subtype’s unique component (*uDep*), we fit a follow-up model, where we flip the order of the Cholesky decomposition model. The resulting model deconstructs each MD subtype into two components: *(i) bDep*, containing all the subtype signal in addition to any signal shared with general MD, and *(ii) rDep*, containing the residual variability in general MD unique from each sub-presentation. Each subtype’s *bDep* (“boosted”) component leverages the power from the general MD construct to identify novel subtype-related biology.

### Stratified Genomic SEM

Stratified Genomic SEM^30^ was applied to identify functional annotations disproportionately enriched for latent MD subtype components. Functional annotations refer to a set of genetic variants that exhibit shared functional characteristics, tissue expression, or synergistic action. We first applied multivariable stratified LDSC (S-LDSC) to obtain a set of stratified genetic covariance and sampling covariance matrices within each annotation. We utilized 118 annotations, including the BaselineLD v2.2^32^ and tissue-informed annotations from GTEx^33^, DEPICT^34^, and Roadmap^35^. Next, we used Genomic SEM’s *enrich* function to estimate functional annotations disproportionally enriched for the MD subtype’s latent components. We applied an FDR-corrected significance threshold for the enrichment of each subtype’s analyzed latent component.

### Transcriptome-Wide SEM

We utilized Transcriptome-wide SEM^30^ (T-SEM) to identify tissue-specific gene expression for latent components of each MD subtype. We first applied FUSION^36^ to perform univariate, summary-based TWAS for each MD subtype and the PGC Freeze 3 MD GWAS. 14 sets of publicly available functional weights were utilized from the GTEx v8^33^, including functional weights for 13 brain tissue types, and PsychEncode^37^, including functional weights for the prefrontal cortex. In total, this yielded 71,331 imputed genes across the different tissues. As the same gene is often present across multiple tissues, this reflects 19,281 unique genes. We then input the univariate FUSION statistics into Genomic SEM’s *read_fusion* function, which rescales the output from FUSION into gene expression-phenotype covariances, standardized with respect to the phenotypic variance. This step harmonizes gene expression estimates to be on the same scale as estimates derived from LDSC. We then estimated T-SEM in Genomic SEM to evaluate the tissue-specific gene expression for each MD subtype’s latent components.

## Results

### GWAS of Child-Onset MDD

We conducted the first publicly available GWAS of child-onset major depression (MD) in the UK Biobank (UKB) across 61,346 MHQ-screened participants (7,965 cases meeting broad MD criteria at ≤18 years of age and 53,381 ancestry-matched controls). We uncovered two independent genome-wide significant risk loci physically proximal to five genes (see Fig. 1, Supplementary Table 2), with the LDSC intercept (1.0), indicating minimal confounding.

**Figure 1.**
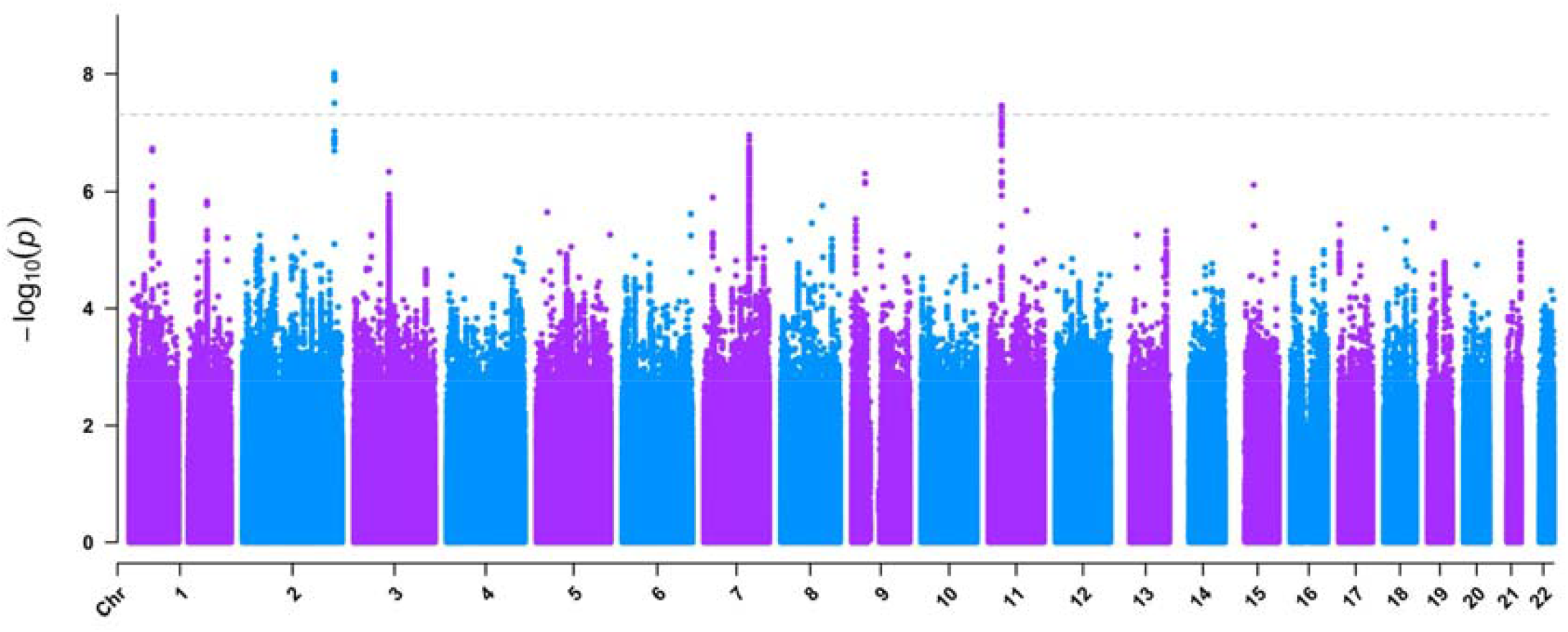
Manhattan Plot for Child-onset MD *Note*. The chromosomes are displayed on the x-axis, and the −log10 *p*-values for the association between each SNP and the physical illness factor are displayed on the y-axis. The dashed gray line denotes the two-sided genome-wide significance threshold of 5×10−8.

### MD Subtypes are Genetically Unique from General MD

We estimated genome-wide genetic correlations (*r*_*g*_) in LDSC to quantify the proportion of genome-wide common variant risk overlap between subtypes and general MD. Perhaps most surprising, we observed only a small genetic correlation between the two MD subtypes (treatment-resistant and child-onset MD *r*_*g*_ = 0.27, *SE =* 0.11). Additionally, we find modest relationships between each subtype and general MD (child-onset *r*_*g*_ = 0.67, *SE =* 0.04; treatment-resistant *r*_*g*_ = 0.58, *SE =* 0.06). We then applied Genomic SEM to decompose the genetic architecture of each subtype into a component shared with general MD and a unique component (see Fig. 2A). This decomposition revealed that most of the genetic signal for these two subtypes was distinct from general MD, with 55% (*SE =* 12%) of the total genetic variance in child-onset and 66% (*SE =* 21%) of the signal in treatment-resistant MD unique from general MD (see Fig. 2B). These findings provide direct, genome-wide evidence supporting the heterogeneity dilution hypothesis, indicating that over half the genetic architecture of these clinically significant subtypes, traditionally lumped in conventional GWAS of MD, is distinct from general MD.

**Figure 2.**
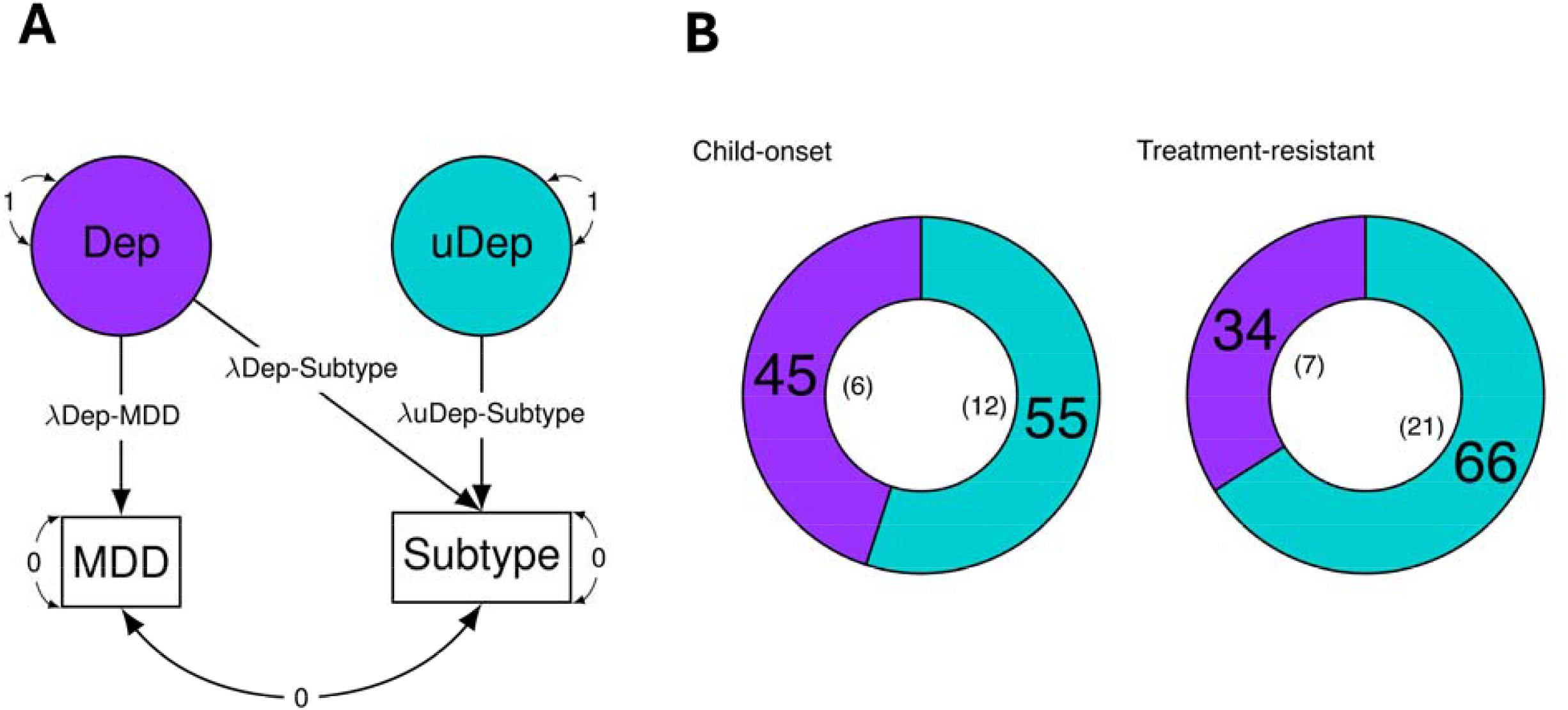
Isolating Genetic Signal Unique to MD subtypes *Note*. (*A*) Single-headed arrows represent regression paths, and curved double-headed arrows represent residual genetic variance components. *Dep* represents a latent factor containing signal from the most recent PGC MD GWAS and whatever is shared with each subtype. *uDep* represents a latent factor capturing variance in each subtype orthogonal to (e.g., unique from) the general MD construct. (*B*) Percentage of variance in each subtype shared with the general MD construct, displayed in purple, and unique from the general MD construct, displayed in blue. The corresponding standard errors are in parentheses inside each circle.

### Subtype-Specific Genetic Overlap with Clinically Relevant Traits

Providing characterization for these subtypes’ latent genetic components, we examined *r*_*g*_ with a panel of clinically relevant external traits (see Supplementary Tables 3-6). We further compare the pattern of genetic correlations exhibited by each subtype’s unique component, *uDep*, to the pattern of *r*_*g*_*’s* exhibited by general MD, as well as the total subtype signal (see Fig. 3).

**Figure 3.**
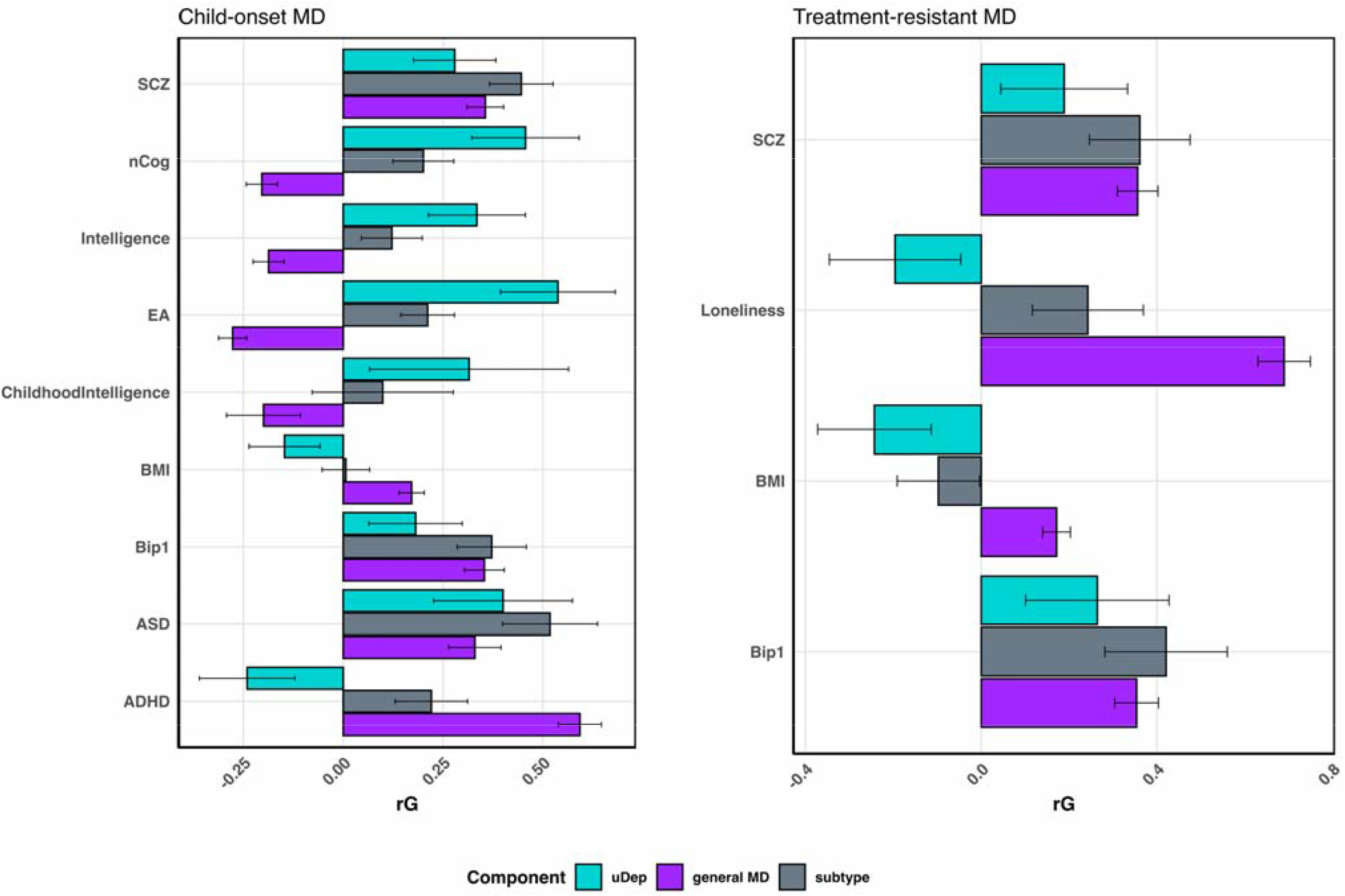
Genetic Correlations with Subtype-Specific Signal *Note*. All presented genetic correlations surpassed an FDR-corrected significance threshold. The light blue bar depicts the unique signal in each subtype (*uDep*), the gray bar depicts the total signal from the subtype, and the purple bar depicts general MD from the most recent PGC GWAS. Error bars depict +/−□95% confidence intervals. SCZ: schizophrenia; nCog: non-cognitive residuals of educational attainment; EA: educational attainment; BMI: body mass index; Bip1: bipolar disorder type I; ASD: autism spectrum disorder; ADHD: attention-deficit/hyperactivity disorder.

We observed a distinct pattern of genetic overlap for the unique component (*uDep*) of child-onset. Notably, this included moderate genetic correlations between child-onset MD’s *uDep* component and educational attainment (EA: *r*_*g*_ = 0.54, *SE =* 0.07) and intelligence (*r*_*g*_ = 0.33, *SE =* 0.06), a pattern observed in the opposite direction for the general MD construct (EA: *r*_*g*_ = -0.28, *SE =* 0.02; intelligence: *r*_*g*_ = -0.19, *SE =* 0.02). Interestingly, amongst cognitive and achievement measures, one of the strongest effects observed with child-onset *uDep* was with the non-cognitive residual of educational attainment (*r*_*g*_ = 0.46, *SE =* 0.07), implicating residual factors facilitating education-facilitating behavior orthogonal to cognition. Consistent with a developmental profile, child-onset *uDep* also exhibited positive common variant risk overlap with childhood intelligence (*r*_*g*_ = 0.32, *SE =* 0.07) and autism spectrum disorder (*r*_*g*_ = 0.40, *SE =* 0.09), aligning child-onset’s unique liability with early-emerging cognitive/behavioral traits and neurodevelopmental features.

Similar divergent associations were observed for treatment-resistant MD’s *uDep* component. Interestingly, we find a negative association between the unique genetic component of treatment-resistant MD and BMI (*r*_*g*_ = -0.24, *SE =* 0.07), observed in the opposite direction for general MD (BMI: *r*_*g*_ = 0.17, *SE =* 0.02), suggesting a unique pattern of symptom directions which may confer risk for treatment-resistant MD (e.g., weight loss). Similarly, loneliness exhibited a negative association with *uDep* (*r*_*g*_ = -0.20, *SE =* 0.08), which reflects the fact that the overall signal for treatment-resistant MD was less positively correlated with loneliness (*r*_*g*_ = 0.24, *SE =* 0.06) than general MD (*r*_*g*_ = 0.69, *SE =* 0.03). Further, we find modest but significant genetic correlations between treatment-resistant *uDep*, schizophrenia (SCZ: *r*_*g*_ = 0.19, *SE =* 0.07), and bipolar disorder type 1 (BD-I: *r*_*g*_ = 0.26, *SE =* 0.08). Interestingly, we observed an insignificant relationship between treatment-resistant *uDep* and bipolar disorder type II (BD-II: *r*_*g*_ = 0.08, *SE =* 0.14), despite positive *r*_*g*_*s* with general MD (*r*_*g*_ = 0.70, *SE =* 0.05) and the full subtype signal (*r*_*g*_ = 0.47, *SE =* 0.12). The relationships between treatment-resistant *uDep* and BD-I and SCZ were smaller than the *r*_*g*_*s* between the total treatment-resistant signal and each of these disorders (SCZ: *r*_*g*_ = 0.36, *SE =* 0.06; BD-I: *r*_*g*_ = 0.42, *SE =* 0.07). This indicates that liability for thought disorders contained within treatment-resistant MD includes both subtype-specific shared risks on top of shared pathways with general MD. Together, these findings suggest that the variance unique to treatment resistance overlaps with genetic variation associated with lower BMI and increased risk for thought disorders (e.g., SCZ and BD-I).

### Leveraging Shared Signal with General MD Reveals Novel Subtype Pathways

We applied Stratified Genomic SEM to investigate patterns of functional enrichment associated with child-onset and treatment-resistant MD. We found no significant *uDep* annotation for either subtype, with treatment-resistant MD also exhibiting no significant annotations in the reversed model that increased power to examine total treatment-resistant signal (*bDep*) by pulling in shared signal with the better powered general MD GWAS (Fig. 4A). Analysis of child-onset MD’s *bDep* component revealed significant enrichment in nine functional annotations (Fig 4B). Notably, this included one novel association (Weak Enhancer^38^, *p =* 1.26 E-3) relative to the subtype itself and general MD (see Supplementary Table 7). The fact that this annotation was not identified for the better-powered general MD GWAS tentatively indicates that the regulatory signal in this weak enhancer annotation is more relevant to child-onset MD. We further identify eight significant annotations for child-onset MD’s *bDep* component that were also identified in the 91 FDR significant annotations for general MD. This included active enhancer marks (H3K27ac) in the Anterior Caudate (*p =* 1.24 E-3), Cingulate Gyrus (*p =* 1.79 E-3), and Dorsolateral Prefrontal Cortex (*p =* 3.91 E-3), as well as primed/active enhancer marks (H3K4me1) in the Angular Gyrus (*p =* 8.80 E-4). For these eight annotations also identified for general MD, this indicates that the shared signal between this subtype and general MD includes genetic variation impacting regulatory elements active in the brain.

**Figure 4.**
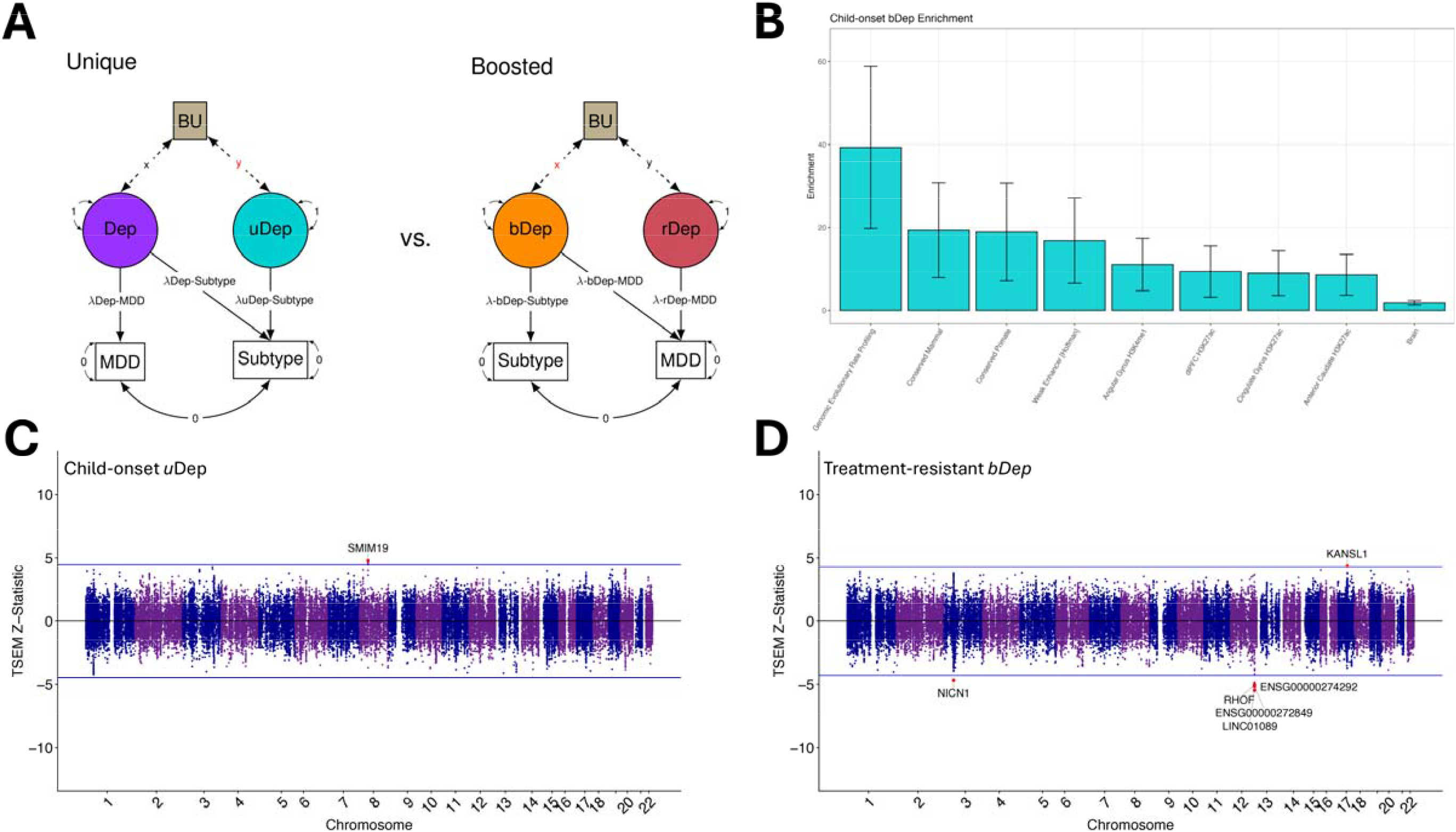
Biological Annotation of MD Subtypes *Note*. (*A*) Schematic of the Cholesky decomposition models applied to biologically annotate latent MD subtype components. Single-headed arrows represent regression paths, and curved double-headed arrows represent residual genetic variance components. BU refers to the biological unit (functional enrichment, tissue-specific gene expression) tested for association with each latent component. (*B*) Stratified Genomic SEM results for child-onset MD’s *bDep* component. All annotations surpassed an FDR-corrected significance threshold. Bars represent the point estimates with error bars depicting 95% confidence intervals. (*C*) Miami plot of gene expression results for unique signal (*uDEP*) in child-onset MD. (*D*) Miami plot of gene expression results for treatment-resistant MD’s *bDep* component. For Panels *C* and *D*, the top and bottom blue lines represent the FDR-corrected significance threshold. Positive versus negative values in both plots indicate upward versus downward patterns of associated gene expression, respectively.

Applying Transcriptome-wide SEM, we uncovered significant tissue-specific gene expression for child-onset MD’s *uDep* component. We found that upregulation of *SMIM19* was robustly associated with child-onset *uDep* across thirteen brain tissues (see Fig. 4C; Supplementary Table 8). Although upregulated *SMIM19* gene expression was also significantly associated with general MD, we find that it is more robustly associated with child-onset’s *uDep* component, with expression of this gene in the bottom 4% of significantly associated genes for general MD. We identified no significant tissue-specific gene expression for treatment-resistant *uDep*. We went on to estimate gene expression for treatment-resistant MD in the power boosted model (*bDep*), finding significant associations with upregulation of *KANSL1* and downregulation of *NICN1, RHOF*, and *LINC01089* (see Fig. 4D; Supplementary Table 9). These genes were also identified in univariate TWAS for this subtype and general MD. While these genes were not novel, their emergence as top hits for this subtype stands in contrast to the fact that they were comparatively much farther down the significance list for general MD. For example, expression of *KANSL1* was among the weakest 5% of significant signal for general MD. Across both subtypes, these may be genes with particular relevance to treatment-resistant or childhood-onset risk pathways and correspondingly reflect putative targets for future mechanistic investigation into divergent etiological pathways within MD.

## Discussion

Using Genomic SEM, we partitioned the common variant architecture of MD to test whether the general diagnostic label masks etiologically distinct subtypes. We find strong genome-wide support for a heterogeneity dilution hypothesis, with 55% and 66% of the common variant genetic signal in child-onset and treatment-resistant MD found to be unique from a conventional GWAS of general MD. Subtype-specific signal also exhibited strikingly divergent patterns of genetic overlap with clinically relevant outcomes. For child-onset MD, this included shared risk with early-emerging cognitive and behavioral traits and selective neurodevelopmental features. For treatment-resistant MD, its unique signal overlaps with genetic variation associated with decreased BMI and loneliness and increased risk for SCZ and BD-I. This profile of clinical correlations closely aligns with how others have described what differentiates melancholia as a syndrome unique from MD^39^, including metabolic chateristics^40,41^, diminished social reactivity, and increased liability for disorders with psychotic features (e.g., SCZ and BD-I).

Biological annotation revealed additional pathways that may be uniquely relevant to these subtypes: transcriptomic and functional enrichments that were at the bottom percentiles of significant signal in general MD were among the most robust associations for the subtypes. At the transcriptome-wide level, gene-expression signals that emerge among the strongest associations for the child-onset MD *uDep* component appear substantially weaker in transcriptome-wide analyses of general MD. More broadly, the ability to identify robustly associated gene expression for sub-presentations of MD, present at the lower ends of significance for the general disorder, highlights the framework’s capability to isolate subtype-specific genetic signals when underpowered for biological annotation. This is most clearly illustrated by stratified genomic SEM of child-onset MD, which revealed novel functional enrichment for child-onset MD *bDep* relative to the general diagnostic construct. These findings across genome-wide, functional, and transcriptomic analyses collectively indicate that a substantial fraction of MD liability resides in subtype-specific components with distinct pleiotropic and biological signatures, which have direct implications for nosology and future molecular genetic investigations of MD.

The formulation of MD as a diagnostic construct has undergone substantial shifts over the 20^th^ century^42,43^. Early editions of the Diagnostic and Statistical Manual of Mental Disorders (DSM) described several depressive syndromes that were later collapsed into a unitary MD framework. The third edition of the DSM emphasized this consolidation, motivated in part by concerns about the poor inter-rater reliability of theoretically derived subtypes and by enthusiasm for pharmacological treatments that appeared to act on a common depressive syndrome. Symptom-based criteria and duration thresholds improved diagnostic agreement. However, contemporary large-scale intervention studies and molecular genetic investigations have established that the MD diagnostic umbrella still contains substantial heterogeneity^18^ and that broad, minimally phenotyped definitions have important limitations for biological discovery^44^. Consequently, there is now potential to use increasingly available biological data to inform nosological approaches, rather than treating nosology as fixed and our biological investigations as downstream.

Evidence in favor of the heterogeneity dilution hypothesis in this study carries both nosological and statistical implications. The variance captured by subtype-specific components in child-onset and treatment-resistant MD indicates that conventional GWAS of general MD are likely estimating an effect averaged over multiple, partially distinct liabilities, attenuating subtype-specific effects, and resulting in a diffuse MD signal. Despite both being presentations of MD, child-onset and treatment-resistant MD share only modest common-variant liability (*r*_*g*_ = 0.27, *SE* = 0.11). This degree of overlap is substantially lower than that observed between subtypes described in our current nosology, such as bipolar I and bipolar II disorder^17,45^ (*r*_*g*_ = 0.83), and more closely approximates estimates of separate diagnostic constructs, such as schizophrenia and general MD^15^ (*r*_*g*_ = 0.33). From a common-variant perspective, these two forms of MD appear, therefore, as genetically distinct disorders rather than minor variations on the same liability continuum. Our results provide a genetic context for a long-standing literature^46^, suggesting that current MD criteria may be lumping etiologically distinct entities under a single diagnostic label.

### Limitations and Future Directions

We note several limitations in this study, primarily the restriction to European-like genetic ancestry samples due to limited data availability and methods for fitting structural equation models across other genetic ancestries. We hope that as datasets increase in size over the next several years, extensions to this work will be developed to improve the representativeness of our findings. Another limitation comes from the retrospective reporting of the age of onset for major depression in the UK Biobank (UKB). In addition, given the limited number of clinically assessed MD cases in the UKB, we rely on a broad MD definition^18^ in addition to the subtype specifier. We hope that as MD subtype data become more widely available, deeper phenotyping approaches^44^ can be applied to gain further insight into the etiology of child-onset MD. We also note that our conclusions should be interpreted as specifically reflecting levels of common-variant risk sharing (SNPs with a MAF > 1%). Future work should seek to evaluate the level of biological distinctiveness in the context of local risk sharing^47^ and rare-variant liability.

### Conclusion

Leveraging Genomic SEM to decompose the common-variant genetic architecture of major depression relative to subtypes therein, we demonstrate that child-onset and treatment-resistant MD contain substantial subtype-specific liability obscured when MD is investigated as a unitary outcome. We find that more than half the common-variant signal for each subtype is unique from the most recent PGC GWAS of general MD. Collectively, these results provide critical context for an evolving nosological dilemma in MD: balancing the risks of indiscriminate lumping against those of excessive splitting. More specifically, our findings challenge the practice of lumping all MD presentations as interchangeable manifestations of a biologically homogenous disorder. Alternatively, they support a genetically informed approach where subtypes are split based on biological evidence for their distinctiveness. At the same time, we also argue against a return to theory-driven proliferation of depressive subtypes and instead support a disciplined strategy in which subtypes are split when they demonstrate divergent biological validity. More broadly, our findings illustrate how molecular genetic investigations can begin to inform outstanding nosological questions, rather than serving to biologically reify existing nosology.

## Data Availability

All data produced are available online at https://doi.org/10.6084/m9.figshare.31329133.

## Data Availability

No new data were collected for this project and all data utilized in this study were from publicly available sources. The data sources for each of the GWAS summary statistics are available in Supplementary Table 3. GWAS summary statistics for child-onset MD are available at https://doi.org/10.6084/m9.figshare.31329133.

## Code Availability

Genomic SEM is publicly available software and can be accessed at https://github.com/GenomicSEM/GenomicSEM.

## Acknowledgments

JML and LSS are supported by NIMH Grant T32MH016880. ADG is supported by NIA Grant R01AG073593. ADG and AS are additionally supported by NIA Grant U01AG083829.

## Notes

### Competing Interest Statement

The authors have declared no competing interest.

### Author Declarations

This study utilized data from the UK Biobank and publicly available data from the Psychiatric Genetics Consortium.

